# Identifying the role of household immunity in driving individual dengue virus infection risk

**DOI:** 10.1101/2023.02.24.23286422

**Authors:** Marco Hamins-Puértolas, Darunee Buddhari, Henrik Salje, Derek A.T. Cummings, Stefan Fernandez, Aaron Farmer, Surachai Kaewhiran, Direk Khampaen, Sopon Iamsirithaworn, Stephen J. Thomas, Timothy Endy, Anon Srikiatkhachorn, Alan L. Rothman, Isabel Rodriguez-Barraquer, Kathryn B. Anderson

**Affiliations:** Department of Medicine, University of California, San Francisco, San Francisco, USA; Department of Virology, Armed Forces Research Institute of Medical Sciences, Thailand; Department of Genetics, University of Cambridge, UK; Department of Biology, University of Florida, USA; Emerging Pathogens Institute, University of Florida, USA; Ministry of Public Health, Tiwanond, Nonthaburi, Thailand; Coalition for Epidemic Preparedness Innovations (CEPI), Washington DC, USA; Institute for Immunology and Informatics, Department of Cell and Molecular Biology, University of Rhode Island, Providence, RI 02903; Faculty of Medicine, King Mongkut’s Institute of Technology Ladkrabang, Bangkok, Thailand; Department of Microbiology and Immunology, SUNY Upstate Medical University, Syracuse, NY, USA; Institute for Global Health and Translational Sciences, SUNY Upstate Medical University, Syracuse, NY, USA

**Author notes:** Indicates shared senior authorship (Isabel Rodriguez-Barraquer, Kathryn B. Anderson).

## Abstract

Dengue virus (DENV) infection risk is known to vary substantially, even across small communities, with infections in and around the home driving transmission. However, It remains unclear how the immune status of an individual or household dictate this risk in part due to transmission being dominated by subclinical infections. In this study, we used demographic, household characteristic, and serological data from a multigenerational cohort study of 2860 individuals from 470 households in Kamphaeng Phet, Thailand, to determine the incidence and risk factors for DENV infections. We used hemagglutination inhibition (HAI) antibody titers measured in sequential serum samples to identify subclinical infections through a gradient boosted regression model. This approach identified ∼10% more cases than commonly used methods with approximately 90% of all infections being subclinical. As expected, we found that having higher DENV antibody titers was protective against infection. Individuals were additionally protected if other household members had higher titers suggesting that there are indirect effects of household immunity on the individuals found within a household. Our study provides a framework for inferring subclinical infections and characterizing the epidemiology of DENV infection in households.

## Introduction

Dengue virus (DENV) presents a major public health burden. As many as 105 million individuals are estimated to be infected with DENV annually ^1,2^, primarily in tropical and sub-tropical regions of the world. There is ample evidence that a significant burden of DENV transmission occurs within and around the home, however, individuals residing in neighboring households may experience persistent differences in infection risk ^3^. The drivers of such heterogeneity in infection risk across households remain unknown. Immunity or susceptibility of household members may impact how likely it is that an infection will be brought closer to the home and in turn risk. However, exploring the role of immunity in household transmission is complicated. Firstly the vast majority of infections are subclinical and thus missed by most surveillance systems ^4^. Studies characterizing risk factors for DENV infection are usually biased towards studying infections causing symptomatic disease and not the greater population of infected individuals. In addition, the burden of DENV has historically been concentrated in children and most studies have focused on understanding infection dynamics in this subpopulation. This has led to large gaps in knowledge about risk factors for DENV infection in adults and full households. Recent shifts in the age distribution of dengue cases towards adults observed in much of South Asia make the aforementioned gap an increasingly pressing issue ^5–8^.

Identifying asymptomatic DENV infections in individuals is challenging, but can be done using data from longitudinal serological testing in cohort studies ^9,10^. Most studies that have used longitudinal serological data to identify subclinical infections have defined infections as a four-fold increase in antibody levels between two samples. However, while there is good support for this cut-point in the context of acute/convalescent samples obtained weeks apart, the performance of this cut-point in identifying infections from samples obtained months or years apart is unclear. Due to antibody waning, the sensitivity of this approach is likely to diminish over time, resulting in underestimates of the true number of infections. Additionally, it is unclear if this method underperforms in individuals with high initial antibody titers. Alternative approaches have reconstructed subclinical infections by fitting full probabilistic models that simultaneously characterize antibody kinetics and infection histories, but this type of method is very data intensive, requiring both large numbers of longitudinal serum samples collected frequently and virologically confirmed infections to estimate antibody kinetics ^11^. Such detailed and prospective data are not commonly available.

This study aims to fill gaps in our understanding of risk factors of DENV infection and disease, by using data from an ongoing longitudinal study in Kamphaeng Phet, Thailand. A key feature of this study is that it enrolled multigenerational households, allowing the risk profiles of children, adults and full households to be studied in parallel ^12^. To identify subclinical infections in the cohort, instead of relying on fixed cut-points, we used a classification algorithm that takes yearly paired antibody titers to determine whether an individual was infected between sampling events. This algorithm was trained using data from individuals with confirmed DENV infections. We then used this imputed dataset to characterize the dynamics of DENV infections in this cohort and to study the association between infection risk and a variety of individual and household factors.

## Methods

### Kamphaeng-Phet family-based cohort study

This study used data from an ongoing family-based longitudinal cohort study in Kamphaeng-Phet, Thailand. Details of the design have been previously described ^13^. Briefly, we enrolled pregnant mothers and their multi-generational households. Per the inclusion criteria, a household was eligible for enrollment if a minimum of three members in addition to the newborn consented/assented to participation; the pregnant woman, another child, and an older adult aged at least 50 years. Active surveillance began with the birth of the newborn, with enrollment specimens for the remainder of the household collected prior to the birth of the newborn. In order to ascertain subclinical infections, serum samples were obtained from all participants roughly annually after enrollment. Acute febrile illness events were detected through a combination of active and passive surveillance strategies. Individuals were instructed to notify study staff if an acute febrile illness event occurred. In addition, participants were contacted by the study team on a weekly basis to determine if any member of the household had been febrile since the last contact. Upon discovery of a febrile episode, an illness investigation was triggered, where acute and convalescent blood samples were obtained from the febrile case. If the illness investigation identified a PCR-confirmed case a household investigation was triggered where acute and convalescent samples were taken for the remaining household members. The convalescent samples were taken at 14 and 28 days after the acute sample collection.

This study was approved by Thailand Ministry of Public Health Ethical Research Committee, Siriraj Ethics Committee on Research Involving Human Subjects, Institutional Review Board for the Protection of Human Subjects State University of New York Upstate Medical University, and Walter Reed Army Institute of Research Institutional Review Board (protocol #2119).

### Laboratory methods

All samples obtained during routine visits were tested using hemagglutination inhibition assays (HAIs) to quantify antibody titers against all four DENV serotypes and Japanese encephalitis virus (JEV)^14^. Additionally, all acute and convalescent samples were tested by HAI for all four DENV serotypes and JEV as well as immunoglobulin M (IgM) and immunoglobulin G (IgG) capture ELISAs for DENV and JEV ^15^. All acute samples also underwent DENV reverse-transcriptase polymerase chain reaction (RT-PCR) ^16^. For the purpose of this analysis, we defined a confirmed DENV infection as any case that is RT-PCR positive for any DENV serotype or where both HAI and ELISA results using the acute and convalescent samples were diagnostic of an infection ^12^.

### Statistical analysis

The purpose of this analysis was to investigate individual and household risk factors for DENV infection in this multigenerational cohort. To do this, we first fit a classification algorithm to the yearly HAI data in order to identify subclinical infections. We then used these imputed infections to investigate individual and household-level drivers of infection.

### Training data

We define a positive and negative person period as follows. A total of 90 confirmed DENV infections were identified through the case investigations. Data from the yearly HAIs surrounding these confirmed DENV infections were defined as the positive person periods. For negative person periods we took the remaining full dataset and removed any interval where an individual had a confirmed DENV infection via the illness investigation, or where individuals had a larger than fourfold increase in any one of their yearly DENV serotype HAIs. We then removed any individuals living in the same household during these aforementioned intervals since DENV transmission is known to be clustered within households. This left 3466 intervals that could potentially be used as negative controls from the available 11131 observed intervals (Figure S1). We randomly sampled a third of these to be added to the training data creating a total of 1246 intervals in our training set. The first interval of sampling for newborns was excluded in this analysis due to limited representation in the serologically supported infections that could provide information on maternal antibody kinetics.

### Prediction model

Using training data described above, we ran a gradient boosted regression using the R package xgboost ^17,18^. Unlike in random forest models where multiple independently trained decision trees are combined to determine the overall likelihood of a model, in gradient boosted regression each decision tree is fit on what the previous trained ensemble of trees have misclassified, allowing for refinement on difficult classification problems. The candidate predictors we used to train this model are listed in Table S3. Variables used to summarize the ratio and difference between pre and post-interval DENV titers across serotypes (maximum, minimum, geometric mean, and variance) were calculated at the individual serotype level and then the summary statistic of interest was quantified across all four serotypes.

### Model fit

For hyperparameter tuning we utilized a random search approach within a nested cross validation approach where we initially split the training data into four cross-validation sets and subsequently performed hyperparameter tuning on each subset using five-fold cross validation. Model performance was quantified using the hold out set. Prior to each random search run we randomly downsampled the dataset to balance the number of positive and negative person periods. We performed this random search approach a total of 5000 times and saved the top 100 performing models evaluated on the held out cross validation set with the lowest log-loss value. The average predicted classification score (bounded between 0 and 1) for these 100 runs was taken to be the probability the individual was infected in that yearly interval. Intervals assigned a value greater than or equal to 0.5 were considered to be DENV infections.

### Predicting subclinical DENV infections

We subsequently fit the models with the lowest log-loss values on the entire training dataset and predicted the presence or absence of infections in the remaining intervals that make up the evaluation data set. We used the training labels as ground truth and subsequently analyzed risk factors for the entire dataset.

### Characterizing risk factors of DENV infection and disease

We fit a series of univariate and adjusted logistic regressions to characterize how DENV infection risk is a function of temporal, individual, and household factors. These models were run using the glmmTMB function found within the glmmTMB package in R ^19,20^. We fit all models using a binomial GLM with a logit link function. All generalized linear models were optimized using the nlminb method found in the stats package. Only household random effects were incorporated into the model as the inclusion of both household and individual random effects led to singular fits.

### Individual and household level risk

We first tested whether demographic factors were associated with risk including age, sex, and employment. We binned individuals into five age bins (1-5, 5-18, 18-30, 30-50, and 50+). Individuals under 1 year were excluded as they will usually have maternal antibodies, which would complicate this analysis due to different kinetics. Sex was defined upon enrollment into the cohort. Further information on individual and household related covariates can be found in the supplemental information.

We subsequently performed analyses to quantify how household composition and infection history impacted risk for an individual. Data on household composition consisted of the number of newborns, individuals between 1 and 5, individuals between 5 and 18, and adults all of which were broken down by sex. We fit models to estimate how the number of individuals in each of these bins impacted infection risk. For the analysis on infection history we took an individual of interest’s household and found the attack rate from the previous interval for all other members. We fit models to assess how the attack rate, the proportion of the household members who were inferred to have an infection, in the previous interval (categorized into three sets defined as containing strictly 0, (0.2), and [0.2,1]) impacted the infection risk. The distribution of these values was zero inflated and skewed right due to many households having no infections in the previous interval. Note that we removed the individual of interest in determining both the household composition and attack rate of the household in order to isolate how the household is impacting risk. For both of these covariates we fit three logistic regression models, a univariate model, a univariate model with random effects, and a multivariate model with random effects.

Since the goal of these models was to characterize the independent effect of the household-level covariates, each of these multivariate models also accounts for the individual’s average pre-interval HAI titer as well as the month and year of sampling as these have been shown to be important predictors of risk. This ensured the individual’s age, titers, and infection history did not impact subsequent calculations. Confounding effects of household related factors were accounted for in adjusted analyses where household random effects were incorporated. Note that as this analysis required at least two consecutive intervals, around 25% of the intervals were not included leaving 6913 intervals.

We then performed logistic regressions to understand how individual and household immunity impact DENV infection risk. We defined individual immunity to be the geometric mean of HAI titers transformed into log2 space. We defined household immunity for a particular individual to be the geometric mean of HAI titers of the household transformed into log2 space with the individual of interest removed from the calculation. HAI cutoffs of 40 and 66 were chosen for the household immunity covariate as these constituted the 33rd and 66th percentiles. Similar to the previous analysis, we fit three logistic regressions for each, a univariate model, a univariate model with random effects, and a multivariate model with random effects. In addition to these random effects and the covariate of interest, each multivariate model also accounts for the month and year of sampling. The household immunity adjusted model also accounted for the individual’s average pre-interval HAI titer.

## Results

### Cohort description

The cohort study contained 3514 individuals within 515 households that have been enrolled since the study began in September 2015. 2868 individuals within 470 households were eligible for inclusion in this analysis as they completed at least one follow-up from enrollment through the cut-off for this analysis in late March 2022. The analyzed dataset contains data on 11131 “yearly” intervals, with an average of 3.90 intervals per enrolled individual (95% CI 1-6). Characteristics of the analyzed intervals are described in Table 1. These intervals were an average of 407.8 days long (95% CI 229 - 642.75) and took place over six sampling periods (Figure 1A). Not all individuals in a household consented/assented to being sampled, with approximately 80% of potential individuals sampled. Over the study period there were 469 index cases that triggered an illness investigation resulting in the identification of 107 infections, 90 of which occurred between paired yearly samples. These 90 infections consist of 62 PCR positive and with the remaining 28 being identified through serological evidence.

**Table 1.** Summary of covariates and their distribution by interval in addition to across infection prediction. Predicted infections are further subdivided into those with symptomatic and subclinical infections.

| Covariate |  | No infection<br>(N=10,082) | Symptomatic<br>infection<br>(N=77) | Subclinical<br>infection<br>(N=972) | Overall<br>(N=11,131) |
| --- | --- | --- | --- | --- | --- |
| Sex | Male | 4192 (41.6%) | 39 (50.6%) | 425 (43.7%) | 4656 (41.8%) |
|  | Female | 5890 (58.4%) | 38 (49.4%) | 547 (56.3%) | 6475 (58.2%) |
| Age<br>(years) | Mean<br>(SD) | 29.6 (22.2) | 14.5 (11.1) | 22.5 (20.8) | 28.9 (22.2) |
|  | Median<br>[Min,<br>Max] | 26.2 [1.00,<br>100] | 12.4 [1.18,<br>57.3] | 14.8 [1.02,<br>88.3] | 25.0 [1.00,100] |
|  | [1,5) | 1696 (16.8%) | 16 (20.8%) | 229 (23.6%) | 1941 (17.4%) |
|  | [6,18) | 2166 (21.5%) | 38 (49.4%) | 310 (31.9%) | 2514 (22.6%) |
|  | [18,30) | 1732 (17.2%) | 17 (22.1%) | 153 (15.7%) | 1902 (17.1%) |
|  | [30,50) | 2121 (21.0%) | 5 (6.5%) | 135 (13.9%) | 2261 (20.3%) |
|  | 50+ | 2367 (23.5%) | 1 (1.3%) | 145 (14.9%) | 2513 (22.6%) |
| JE<br>vaccination<br>in interval | Yes | 453 (4.5%) | 1 (1.3%) | 68 (7.0%) | 522 (4.7%) |
|  | No | 9629 (95.5%) | 76 (98.7%) | 904 (93.0%) | 10,609 (95.3%) |
| Pre interval<br>titer (HAI) | Mean<br>(SD) | 117 (200) | 31.1 (37.0) | 55.8 (83.6) | 111 (193) |
|  | Median<br>[IQR] | 67.3<br>[14.1,134.5] | 14.1 [10,<br>33.6] | 28.3 [10,<br>67.3] | 56.6<br>[14.1,134.5] |

**Figure 1.**
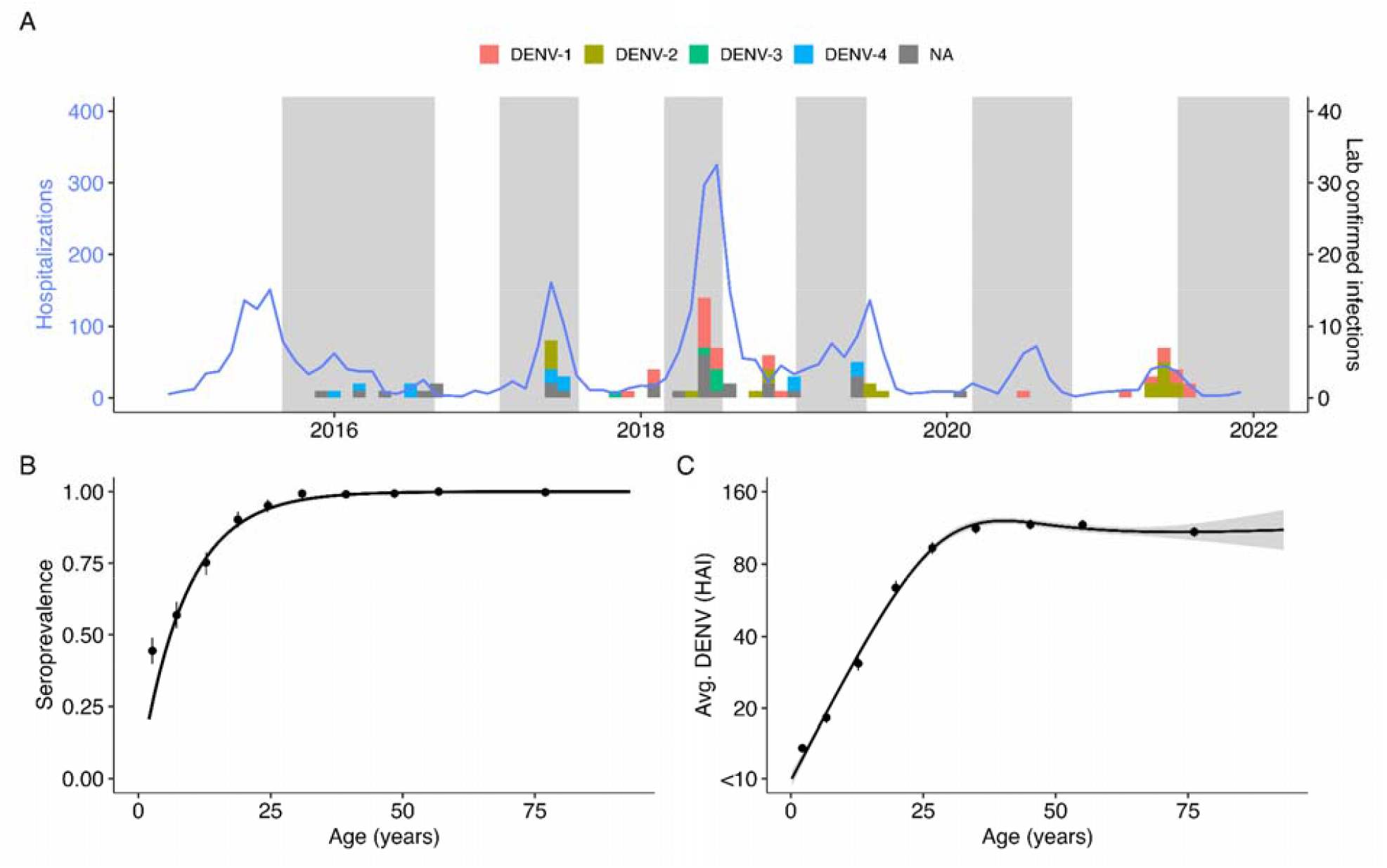
Summary of cohort data. (A) Hospitalization counts for Kamphaeng Phet from 2015-2021 (blue solid line). Bars represent the timing of the confirmed DENV infections used to train the model. Serotype information was ascertained via RT-PCR. Shaded time periods represent active sampling periods during the cohort study when yearly blood draws were taken. (B) Age-stratified seropositive at enrollment for subjects enrolled before 2017. Points are mean seroprevalence found at each tenth percentile age bin. (C) Average DENV HAI titers at enrollment by age. Confidence bounds for B and C are found using a basic nonparametric bootstrap.

### Model performance

We developed a classification algorithm that takes antibody titers measured during yearly visits to infer undetected infections in individuals. Our best fitting model was able to classify our training data with 93.3% sensitivity and 96.6% specificity (Figure S7). The longitudinal design of the cohort study allows for the visualization of HAI trajectories across time for enrolled individuals. Figure 2A illustrates imputed infections for three individuals enrolled in the cohort. The average and maximum ratios of pre to post interval HAI titers are the features of greatest importance for accurate classification defined by the information gain metric (Figure 2B).

**Figure 2.**
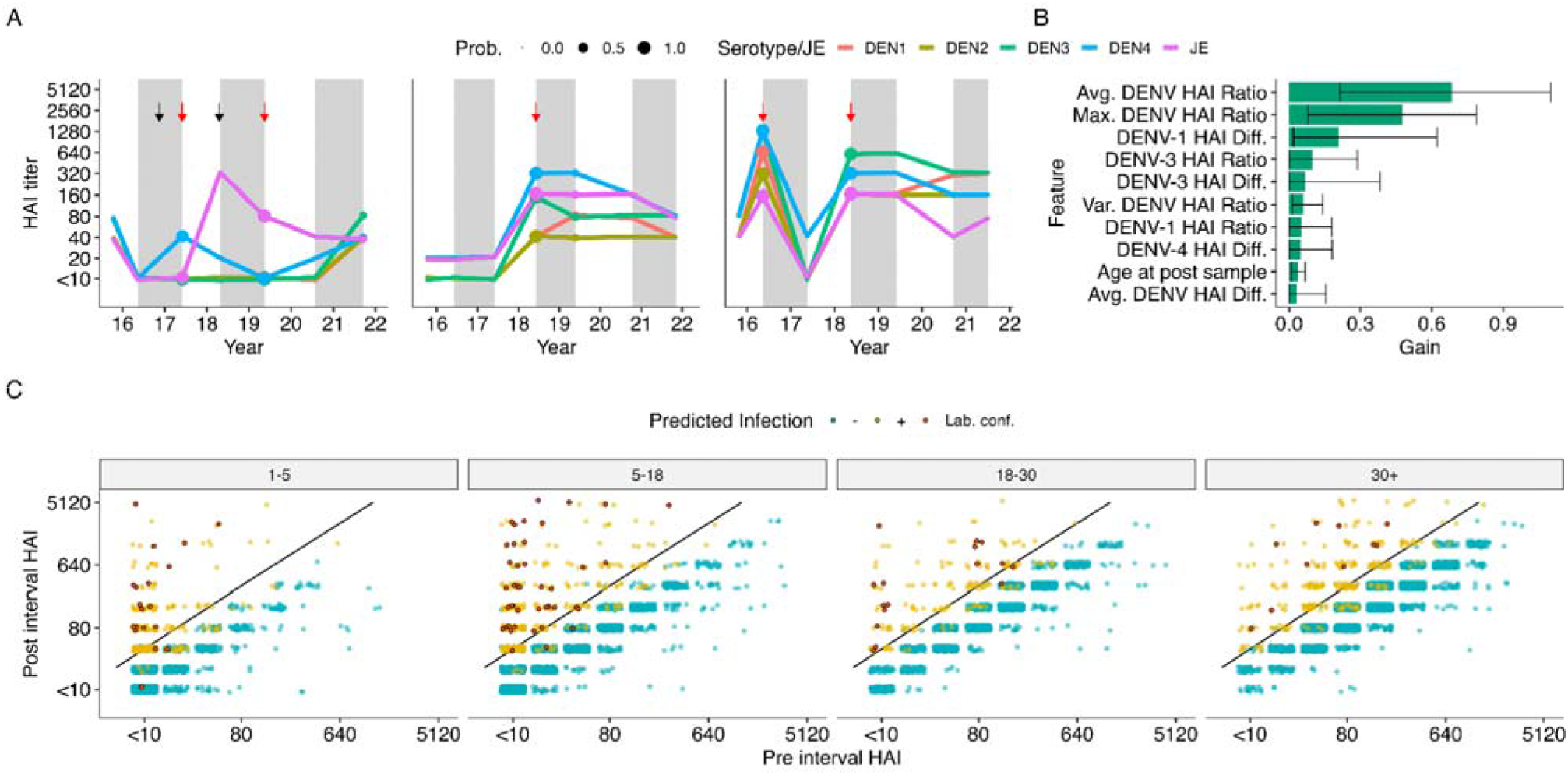
Summary of model performance and fits. (A) Three examples of HAI titer trajectories for all four DENV serotypes and JEV in three subjects. Alternating white and gray time periods represent distinct intervals, separated by the sampled HAIs. The imputed probability of an infection having occurred within an interval is presented by point size at the post-interval sample date while red arrows represent an imputed infection. Black arrows represent JEV vaccination events. (B) Feature importance across 100 model fits. Gain represents the relative contribution of each feature. (C) Pre and post interval HAI titers for the DENV serotype with the largest ratio grouped by age at post-interval sampling event and colored by whether the model predicted a seroconversion. Red points represent the confirmed seroconversions from the training data. A four-fold increase in titers between samples is represented by the black diagonal line.

### Characterizing subclinical infections

Using our best fitting prediction models, we identified 1049 infections in 11131 intervals of observation, or 9.4% of intervals. This translates to 8.42 infections per 100 people per year (95% CI 7.71 - 9.15). This is similar to an estimated annual proportion of seronegative individuals being infected per year of 10.8% derived using a serocatalytic model from age-stratified seroprevalence data (95% CI, 9.9-11.8% using seropositive cutoff as HAI >=20) OR (8.0%, 95% CI, 7.3-8.7% using seropositive cutoff as HAI >=40) (Figure 1B). This force of infection is derived for non-newborns under 30 years old and the methods taken to estimate it are outlined in the supplemental information. We note that the model had high certainty in the assigning of infections for the majority of infections, with 673 of the 1049 intervals with infection being given a probability of greater than 90%. Similarly, the model had high certainty for the absence of infections in the remaining intervals with 8455 of the 10285 intervals being assigned a probability of less than 10% (Figure S8).

We found that the incidence of infections varied by year, with 2018 having higher incidence (Figure 3A). This is the same year where hospitalizations peaked in Kamphaeng Phet across the active years of the study (Figure 1A). Incidence of infection rates peaked amongst school aged children while individuals over the age of 30 all experienced lower incidences (Figure 3B). As expected, the proportion of primary infections (infections occurring in individuals without detectable antibodies to any serotype in any prior visit) was directly related to age, with almost all infections being post-primary (occurring in individuals with HAI antibody titers against at least one serotype greater than 20) after age 25. The ratio of subclinical to symptomatic infections was 13.8:1 (95% CI: 10.0-17.8:1) in the cohort. There was some variability across years and age, with the highest risk of symptomatic disease occurring between the ages of 15-25 (Figure 3D-E). We note that there were only 77 symptomatic infections out of 1049 total infections, leading to wide confidence intervals for these ratios, particularly for years of age-groups with few cases. Out of these 1049 infections, 139 individuals had multiple infection events throughout the study. The probability of having a second or third infection given that the individual had a previous infection peaks between the ages of 10-15, similar to the age range of highest incidence (Figure S2B).

**Figure 3.**
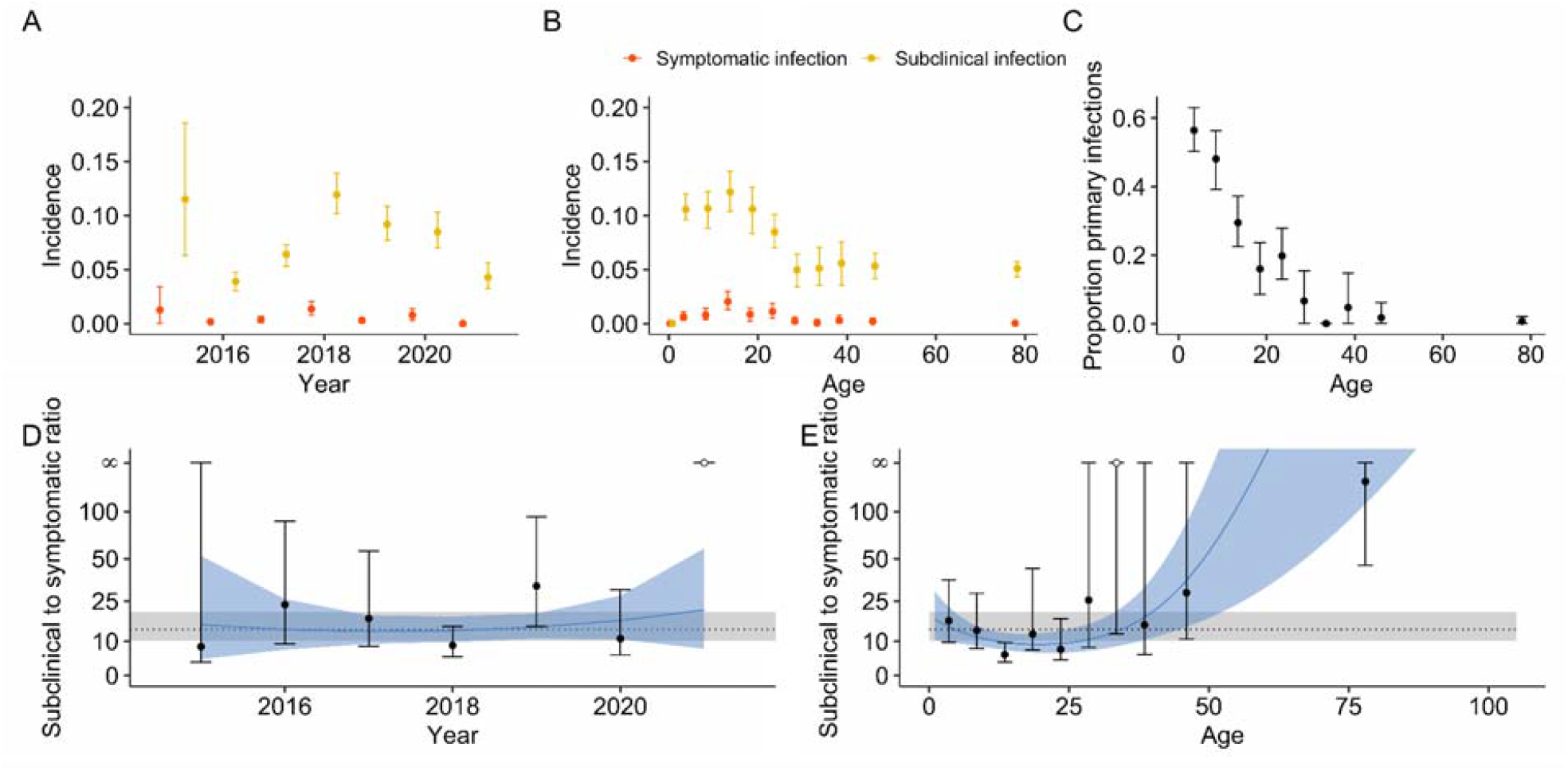
Incidence, proportions of primary infections, and subclinical to symptomatic ratios across time and age. (A-B) Incidence (infections per person-year) for both symptomatic (red) and subclinical (yellow) infections across interval year (A) and age (B). (C) Proportion of primary infections as a function of age. (D-E) ratio of subclinical to symptomatic DENV incidence in the cohort as a function of interval year (D) and age (E). Mean and 95% CIs for the ratio of subclinical to symptomatic DENV incidence are represented by the dotted line and gray regions respectively. Mean and 95% CIs for polynomial fits to time and age are represented by the solid blue line and regions.

### Risk factors for DENV infection

Using the imputed infections from the classification algorithm we investigated which individual and household risk factors were associated with infection risk. When compared to young children between the ages of one and five years old we found that individuals between 5 and 18 as well as those between 18 and 30 were at higher risk of infection with an aOR of 1.44 (95% CI 1.16-1.77) and 1.41 (95% CI 1.06-1.89) respectively. In an unadjusted analysis there was no significant difference in odds of infection by sex when comparing males to females (OR 1.11, 95% CI 0.98 - 1.27). However, our data is consistent with an observed interaction between age and sex in infection risk with women between the ages of 18-40 being at a higher risk of infection than their male counterparts (Figure S4). We also studied how an individual’s occupation may impact their risk and found no differences that attained statistical significance in the adjusted analysis (Table S1).

We subsequently studied how household level factors impact an individual’s risk of infection. No covariates describing the surrounding built environment had a significant impact on dengue risk. However, we did find strong associations between household composition and risk of infection. While the number of individuals living in the household was not associated with risk of infection (aOR 1.00, 95% CI, 0.97 - 1.04), we found that each additional adult in the household reduced the likelihood of infection in the other household members with an aOR of 0.95 (95% CI, 0.90 - 0.99). The presence of each additional newborn and individual between 5 and 18 increased the odds of infection for the other household members with an aOR of 2.13 (95% CI, 1.65 2.75) and 1.09 (95% CI, 1.01 - 1.19) respectively. Although not significant, the presence of each additional individual between one and five increased the odds of infection for household members with an aOR of 1.13 (95% CI, 1.00 - 1.28) (Figure 5A). Analyses stratified by sex revealed a more complex association between household composition and risk. For either sex, each additional newborn increased infection risk for the other individuals living in that household. For older age groups however the associations varied by sex. Each additional male between the ages of 1 and 5 and between 5 and 18 increased risk with an aOR of 1.25 (95% CI, 1.08 - 1.44) and 1.18 (95% CI, 1.06 - 1.31) respectively while additional adult male had no impact on risk. Additional females provided no changes in risk except for adults, where each additional female adult reduced risk with an aOR of 0.88 (95% CI, 0.81 - 0.95) (Figure 5B).

Beyond characterizing the association between household characteristics and composition on dengue risk, we sought to understand the impact of individual and household immunity. Consistent with previous findings, the most important predictor of infection risk during an interval was an individual’s HAI titers at the beginning of the interval (Figure 4). In this analysis, the magnitude of average HAI log2 titers was inversely associated with risks of both symptomatic and subclinical infections. On average, each log2 increase in titers was associated with a 26.3% (95% CI: 23.3 - 29.1%) decrease in risk of infection and a 38.7% (95% CI: 27.9 - 47.9%) decrease in having a symptomatic infection. Interestingly, we also found that household immunity impacted an individual’s risk of infection even when accounting for that individual’s antibody titer. The distribution of these variables is found in Figure S7. Individuals living in households with high immunity (average HAI titers greater than 66) had decreased risk with an aOR of 0.78 (95% CI, 0.63 - 0.96) when compared to those with an average below 40 (Figure 5D). Since household titers are likely to be associated with recent household infection history, we also investigated how household attack rates during a preceding interval (the proportion of individuals within a household that had an imputed DENV infection in the preceding interval) impact future risk. Individuals living in households that had moderate to high attack rates (greater than 20% of household members experiencing an infection) during the previous year were at decreased risk of infection with an aOR of 0.61 (95% aOR CI, 0.49 - 0.77) in comparison to individuals coming from a household with no infections the previous year (Figure 5C).

**Figure 4.**
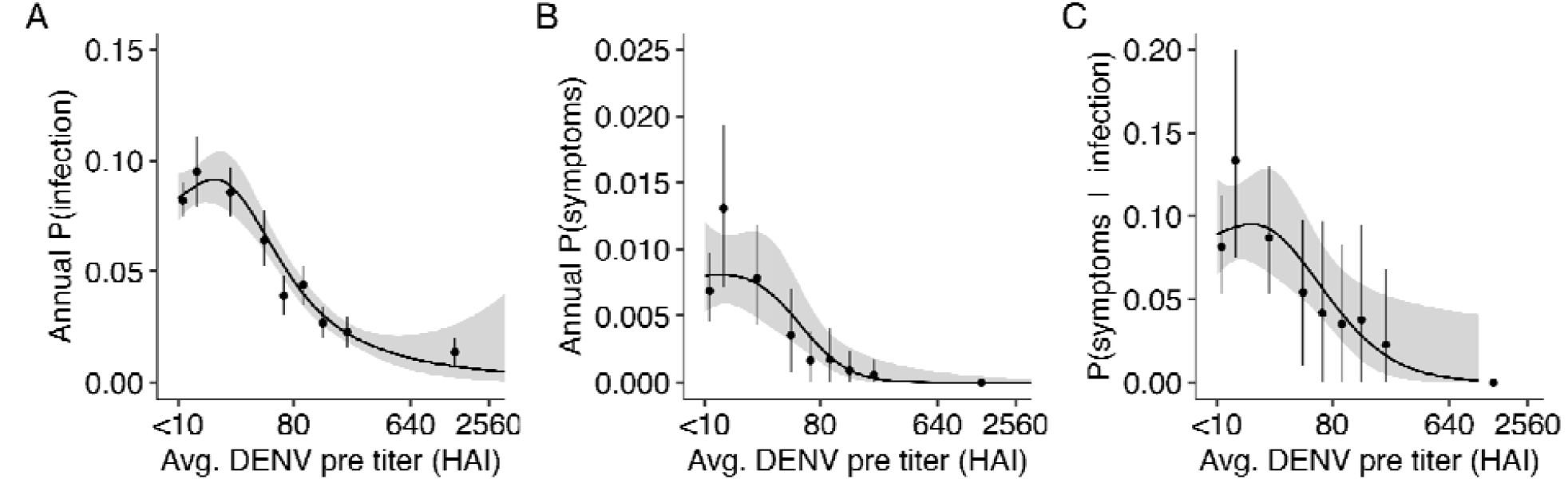
Geometric mean of pre-interval DENV titers across all four serotypes and their impact on probability of infection and symptoms. We first calculated the annualized probability of infection and then fit splines of order three to the data. (A) Probability an individual is infected in a year as a function of their pre-interval DENV titers. (B) Probability an individual is symptomatic in a year as a function of their pre-interval DENV titers. (C) Probability that an individual is symptomatic given that they were infected in the same year as a function of their pre-interval DENV titers.

**Figure 5.**
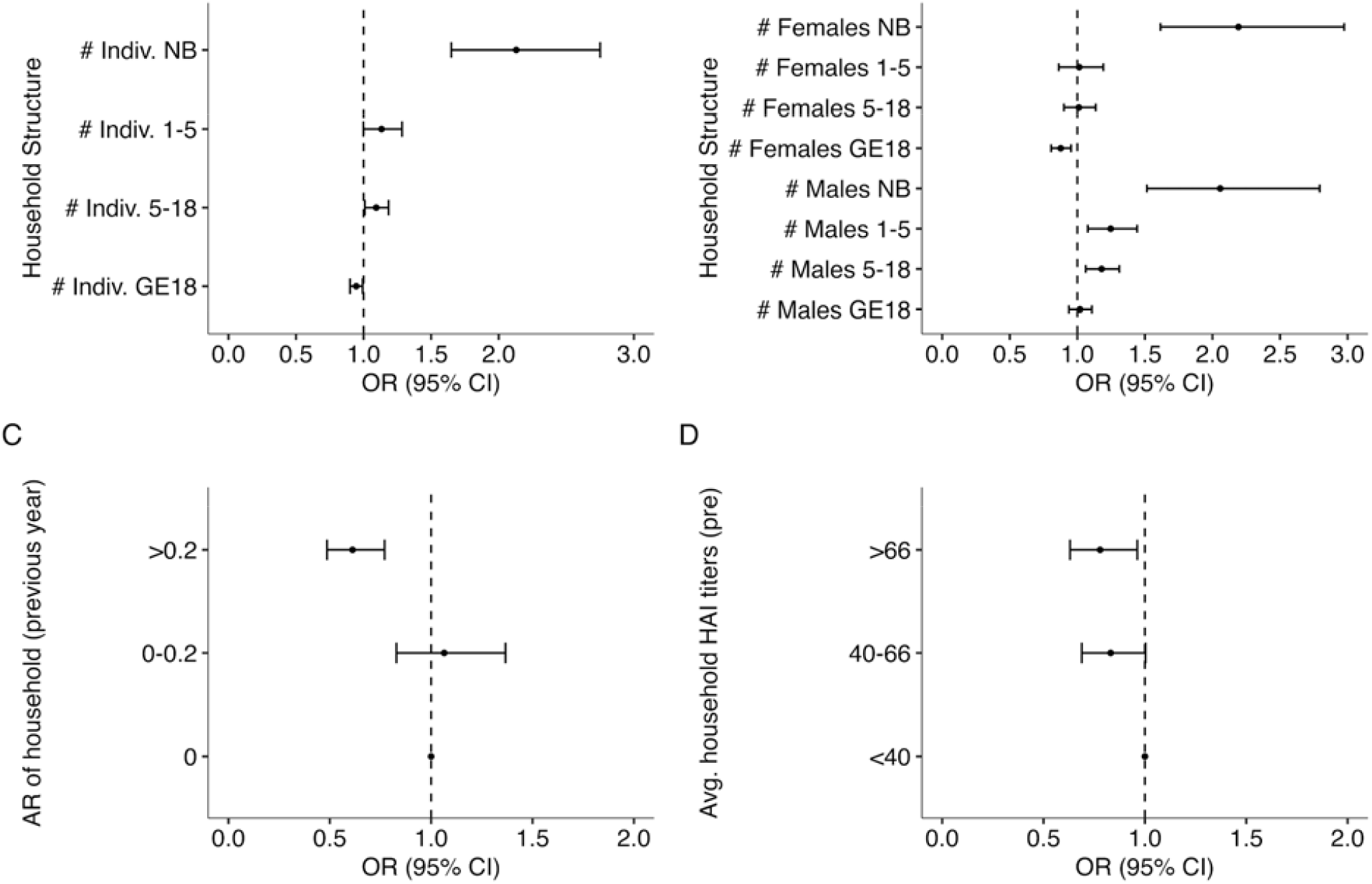
Impact of household composition (A-B), infection history (C) and immunity (D) on risk of infection. (A) Odds ratio for the number of total individuals in various age bins (newborn [NB], from 1-5 years old [LT5], from 5 to 18, and those older than 18 [GE18]) defined at the time of the post-interval sample. (B) Odds ratio for the number of males and females of various age bins (newborn [NB], from 1-5 years old [LT5], from 5 to 18, and those older than 18 [GE18]) defined at the time of the post-interval sample. (C) Previous interval’s attack rate and subsequent odds ratio of infection risk relative to having no infections in the previous interval. (D) Geometric mean of DENV HAI titers for the rest of the household members and subsequent odds ratio of infection risk relative to having an average household HAI titer under 40. All models are adjusted for household random effects, individual pre-interval titers, as well as the year and month of post-interval sample.

## Discussion

In this work we developed a classification algorithm using longitudinal data from a multigenerational cohort in Kamphaeng-Phet, Thailand to reconstruct unobserved DENV infections. This allowed us to investigate specific individual and household level factors that are related to DENV infection risk. Our findings are consistent with a protective effect of higher HAI titers at both the individual and household level. While previous work has demonstrated that higher titers are protective against infection at the individual level, the impact of household immunity and composition on infection risk have not been investigated ^11^. We find that beyond individual titers, there is an independent indirect effect of immunity in other members of the same households.

We studied how several household factors including composition, immunity, and infection history each independently impact risk of infection by DENV. We found that all three independently help determine an individual’s risk. When analyzing household composition we found that each additional adult reduced the likelihood of an infection while each additional young child (1-5 years old) increased the likelihood of infection. Some of this relationship may be explained by the fact that children are more susceptible to infection than their adult counterparts who have already experienced infection and developed immunity in the past. Quite similarly we found that higher levels of household immunity and higher attack rates in the previous year have protective effects on infection risk. All of these factors present a similar picture of household factors, where households with more adults or more recent infections will have more immunity to DENV and in turn alter subsequent infection risk for the members of the household.

At the individual level, our results are consistent with prior studies showing that individual antibody titers are the most important predictor of future DENV infection risks. How this relationship varies across adults is less understood. Here we find that the risk of infection for adults over the age of 30 remains high, at approximately half that of younger individuals. These infections occur in individuals who have been infected two or more times and are in turn multitypically protected. This is particularly relevant to the open question of how long boosting post-infection confers immunity and protection from clinical manifestations. For these same individuals we find a higher subclinical to symptomatic ratio, suggesting that these adults are likely exposed to DENV while simultaneously not experiencing symptoms.

Although the precise process behind our results are likely complex, we hypothesize that they can be mechanistically disentangled into how infections are brought into the home and how they spread within it. In prior work, we hypothesized that shifts in the age structure of people in Thailand have resulted in decreases in the force of infections since highly immune adults live longer and are more likely to have multitypic immunity that can absorb potentially infectious bites from mosquitos that would otherwise bite younger individuals living in the same household ^5,21^. Our results show that the presence of high immunity and recent infection history in the household will provide a form of herd immunity to an individual, regardless of their own immune status. On the other hand, children are more likely to be seronaive and may present a mechanism through which DENV can be introduced into the household. These introductions will subsequently increase the risk that the virus will be transmitted (by mosquitoes) to others in the household, a mechanism that would explain some of the spatial correlation found in this population ^22^. It is interesting to find that household composition, immunity, and infection history have a significant impact on infection risk while covariates associated with the physical structure of the household do not.

This work also provides a framework on which machine learning classification algorithms can be used to predict infection events from yearly serological data. The use of a four-fold rise in titers as a barometer for infection can be useful in some contexts, but we find our approach to be robust and more sensitive to characterize subclinical infections. While previous Bayesian based approaches have been successful at reconstructing dengue infection events ^11,23^, they require substantial temporal information to inform the underlying mechanistic model of antibody kinetics. Our work provides a flexible framework that removes some of the bias of potential model misspecification and instead takes a fully data driven approach to reconstruct infection events.

Our results highlight the importance of multigenerational household studies in order to fully understand the population dynamics of infectious diseases. The protective effects of household immunity had been elusive in prior work, some in the same setting, focusing on children. However, this work also has some limitations. Our model training is limited by the fact that there are only 90 data points used to inform the classification algorithm on how yearly HAI titers can look before and after an infection. If these illness investigations are biased in some manner, this would propagate to our predictions. In particular if primary DENV infections are underrepresented then we may be less capable of catching these primary DENV infections. In addition, development of the training data required that we define individuals who had no infection event during an interval, a difficult task that could further limit this approach. A limitation of the study was the fact that serum samples were taken at yearly intervals. This made it impossible to fully disentangling the timing of infections which would provide important information on how infections propagate in a household. Incorporating additional active sampling events throughout the year in household studies like this one could provide important temporal information to understand this further. Finally, due to study design, most female participants of reproductive age give birth upon enrollment. We are therefore not in a position to examine whether the sex differences found between the ages of 18-40 are due to age or to other biological or behavioral factors (Figure S5) related to pregnancy and giving birth. Further work must be done to fully understand this relationship.

Understanding how immunity impacts the spread of infectious diseases like DENV is a vital question. General principles of epidemiology suggest that individuals with higher immunity are protected from infection and disease, while entire populations can also experience protective effects of population wide immunity. Here we show that this relationship is also found at the household level, something that has not previously been studied in DENV nor other vector borne diseases to our knowledge.

## Data Availability

All data and code associated with the work can be found at https://github.com/marcohamins/role-of-HH-immunity.

## Acknowledgments

We are thankful for all efforts from the data collection team as well as the children and adults involved in the study. The NIH Grant P01 AI034533 and R35 GM138361 supported the work.

## Notes

### Competing Interest Statement

The authors have declared no competing interest.

