## Supplemental information for "Identifying the role of household immunity in driving individual dengue virus infection risk"

*Data and code availability*

All data and code associated with the work can be found at <https://github.com/marcohamins/role-of-HH-immunity>.

*Individual and household related covariate description*

Employment status was updated each interval through a yearly questionnaire. We binned occupations by behavior; all farmers were combined into a single classification, all company employees, government officers, general employees, retail merchants, and unspecified occupation being classified as employed, while students and unemployed individuals remained as their own classifications. We then tested a host of household factors. This included factors associated with potential locations for *Aedes* mosquitoes to breed including the number of water containers, plastic water containers, and plastic bottles all of which we analyzed as log10 counts. In addition, indicator covariates associated with the household being near a source of water and whether the household was supplied water by a pipe were included. Covariates for the physical structure of the house were also included. These include whether the house had a zinc roof, was concrete, had door screens, and the number of toilets outside the home. In addition, whether the house was built on poles, was a single unit, or a townhouse was analyzed along with the garbage management system of the home (car collection versus burnt/buried/dumped).

*Force of infection*

A catalytic model was fit to estimate the force of infection in the population. Baseline seroprevalence of DENV in a subset of individuals enrolled in the study before 2017 were used for this analysis. We fit a model with age only to estimate the general force of infection with a *cloglog* link function ^24^. We ran this model on individuals over the age of one to remove the impact of maternal antibodies. We also removed individuals over the age of 30 as the entire population approaches complete seropositivity around this age. If $\lambda$ is the annual force of infection we are able to translate this to a proportion of susceptible individuals infected per year using the following formula, $1-exp(-\lambda)$.

*Sensitivity Analysis*

We conducted sensitivity analyses on the methods outlined in the *Individual and household level risk* subsection of the methods. The first sensitivity analysis we performed was based on the fact that at times not all individuals in a household were sampled. Those that went unsampled in a household were more likely to be adult males potentially leading to confounding effects of households with more missing data. We in turn perform analyses on all intervals taken from households with more than 80% of their members sampled (Figure S9C-D). The attack rate results held for this sensitivity analysis. We also modeled risk strictly in individuals who are seronaive at the beginning of an interval in order to quantify how these risk factors look in this subpopulation of interest (Figure S10C-D). The analyses for mean household titers demonstrated significant protection for both medium and high immunity when restricting the analysis to these seronaive individuals. In this subpopulation we found no relationship between household attack rate and infection risk.

We ran similar sensitivity analyses on how household composition impacted infection risk. The results obtained when subsetting the data to households with more than 80% of their members sampled and when subsetting to individuals who were seronaive at the start of an interval were similar to one another (Figure S9A-B,S10A-B). Unless noted otherwise we will report both simultaneously. Each additional newborn and individual between the ages of one and five increased the likelihood of infection for other members in the household. Additional adults reduced the odds of being infected, but not significantly so. When stratified by sex we found that each additional female newborn significantly increased the odds of infection while each additional male increased the odds but without statistical significance. Each additional male between the ages of one and five increased the odds of being infected for other members while each additional female had no significant impact. Neither of these sensitivity analyses produced any significant impact of additional adults on infection risk for other household members.

**Table S1.** Odds ratios and adjusted odds ratios for individual and household level covariates of infection risk. Univariate analyses inform which variables are incorporated in the multivariate analyses. Multivariate analyses were conducted using either just household random effects, both household and individual random effects, or household random effects and incorporating individual immunity. Odds ratios and adjusted odds ratios for temporal covariates can be found in Table S2. Abbreviations: aOR, adjusted odds ratio; CI, confidence interval; Nb., number;OR, odds ratio; REF, Reference category.

| **Covariate** | | **Univariate** | **Multivariate**  **House res** | **Multivariate**  **indiv., house res** | **Multivariate house res w/**  **indiv. immunity** |
| --- | --- | --- | --- | --- | --- |
|  |  | **OR**  **(95% CI)** | **aOR**  **(95% CI)** | **aOR**  **(95% CI)** | **aOR**  **(95% CI)** |
| **Individual level** | | | | | |
| Sex | Female | REF | – | – | – |
|  | Male | 1.11 (0.98 - 1.27) | – | – | – |
| Age | [1,5) | REF | REF | REF | REF |
|  | [5,18) | 1.11 (0.93 - 1.33) | 1.16 (0.95 - 1.41) | 1.16 (0.95 - 1.41) | 1.44 (1.16 - 1.77) |
|  | [18,30) | 0.68 (0.55 - 0.84) | 0.74 (0.58 - 0.95) | 0.74 (0.58 - 0.95) | 1.41 (1.06 - 1.89) |
|  | [30,50) | 0.46 (0.37 - 0.57) | 0.41 (0.30 - 0.57) | 0.41 (0.30 - 0.57) | 0.93 (0.63 - 1.35) |
|  | 50+ | 0.43 (0.34 - 0.53) | 0.41 (0.27 - 0.62) | 0.41 (0.27 - 0.62) | 0.88 (0.55 - 1.40) |
| Occupation | Employed | REF | REF | REF | REF |
|  | Farmer | 0.57 (0.27 - 1.19) | 0.68 (0.31 - 1.48) | 0.68 (0.31 - 1.48) | 0.63 (0.29 - 1.38) |
|  | Student | 1.64 (1.17 - 2.29) | 0.78 (0.51 - 1.18) | 0.78 (0.51 - 1.18) | 0.66 (0.43 - 1.01) |
|  | Unemployed | 1.24 (1.02 - 1.50) | 0.82 (0.63 - 1.07) | 0.82 (0.63 - 1.07) | 0.79 (0.60 - 1.04) |
| **Household level** | | | | | |
| Nb. Water container (log10) | | 0.94 (0.78 - 1.14) | – | – |  |
| Nb. Water container - plastic (log10) | | 1.03 (0.90 - 1.18) | – | – | – |
| Nb. Plastic bottles (log10) | | 0.99 (0.86 - 1.14) | – | – | – |
| House type | Poles | REF | – | – | – |
|  | Single | 1.08 (0.91 - 1.28) | – | – | – |
|  | Townhouse | 0.97 (0.67 - 1.40) | – | – | – |
| Garbage management | Burnt / Buried / Dumped | REF | – | – | – |
|  | Car collection | 1.11 (0.97 - 1.26) | – | – | – |
| Concrete house | | 1.08 (0.92 - 1.26) | – | – | – |
| Zinc roof | | 1.00 (0.87 - 1.14) | – | – | – |
| Nearby source of water | | 1.01 (0.88 - 1.16) | – | – | – |
| Water supply by pipe | | 0.91 (0.74 - 1.11) | – | – | – |
| Door screens | | 1.04 (0.96 - 1.13) | – | – | – |
| Number of toilets outside | | 1.03 (0.93 - 1.14) | – | – | – |

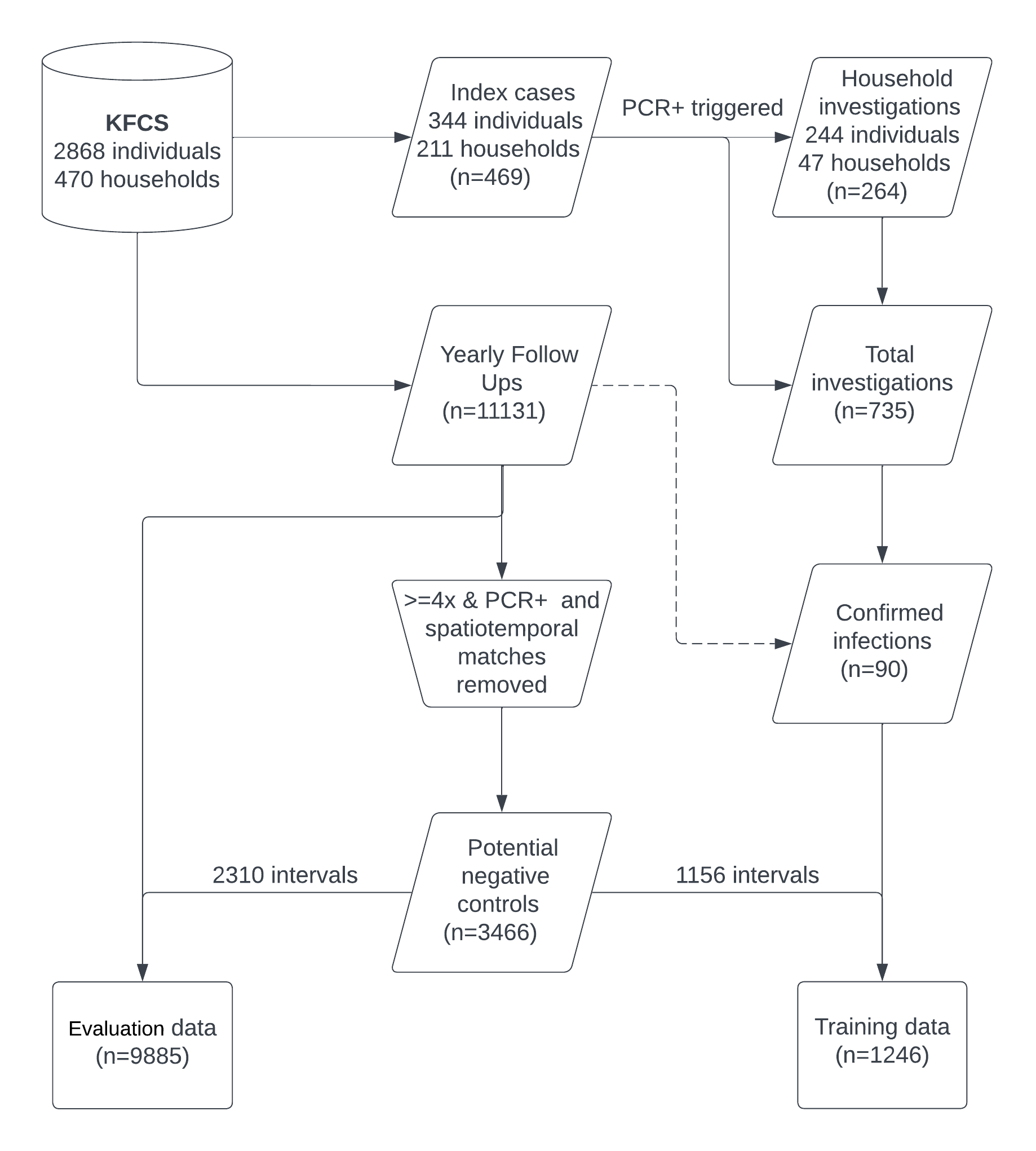

**Figure S1.** Data flow from original stored data to the training and evaluation data sets utilized for the analysis. The number of intervals found at each step is noted by *n*. A total of 470 households and 2868 individuals were enrolled in our cohort. Index cases were tested for DENV infections using PCR. Positive test results led to household investigations for the remaining household members. This led to 735 total intervals of which 90 had a confirmed DENV infection which formed part of our training data. From the complete dataset we then removed these confirmed DENV infections, their spatiotemporal matches, as well as any intervals that had a maximum DENV ratio at or above a four-fold rise. This led to 3466 total potential negative controls from which one third were randomly chosen to be added to the confirmed infections that would make up our training data (n=1246). The remaining intervals were kept in our evaluation data (n=9885).

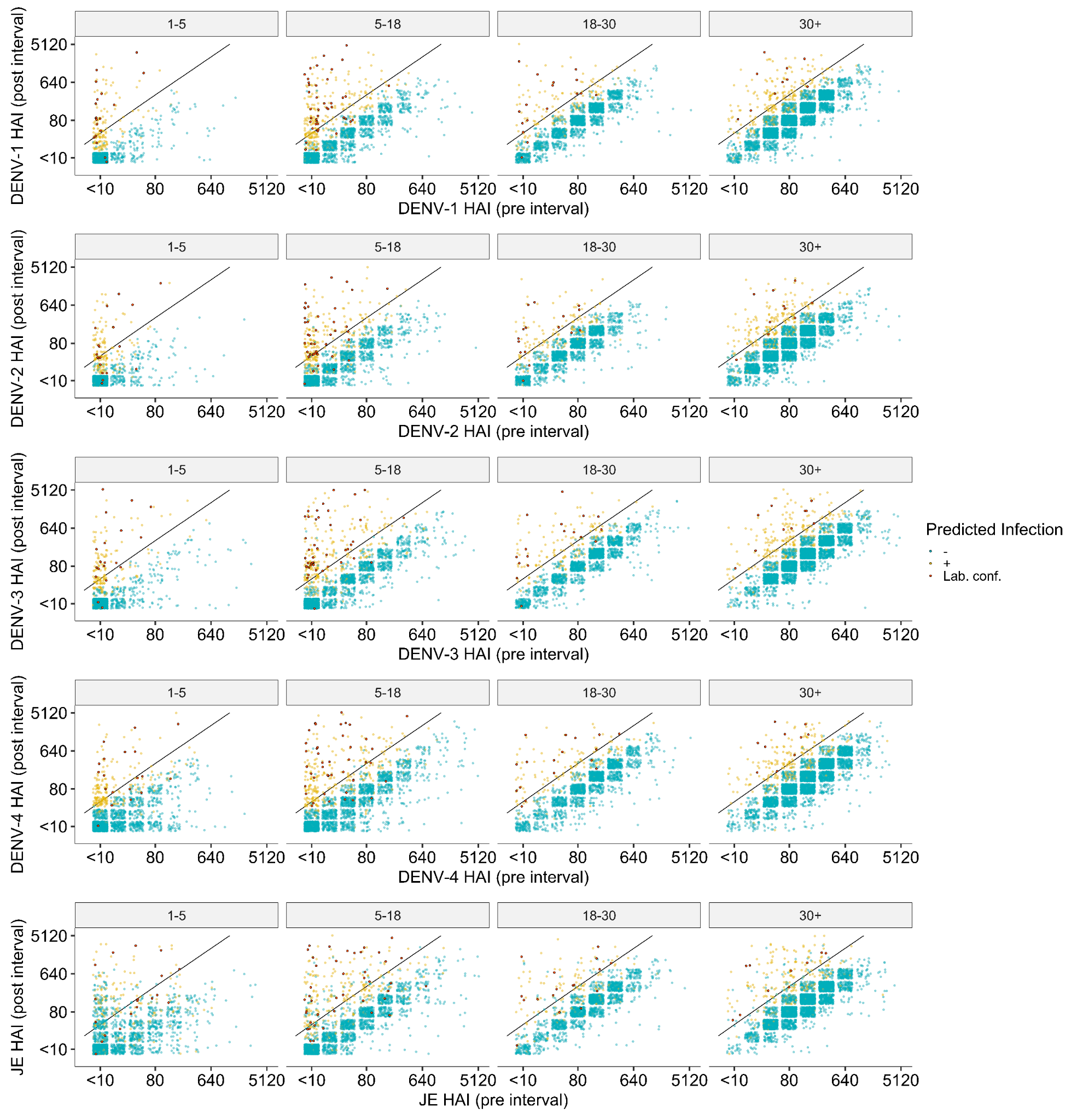

**Figure S2.** Pre and post interval HAI titers for all DENV serotypes and JEV grouped by age at post interval age and colored by whether the model predicted a DENV infection. Red points represent the confirmed seroconversions from the training data. A four-fold increase in titers between samples is represented by the black diagonal line.

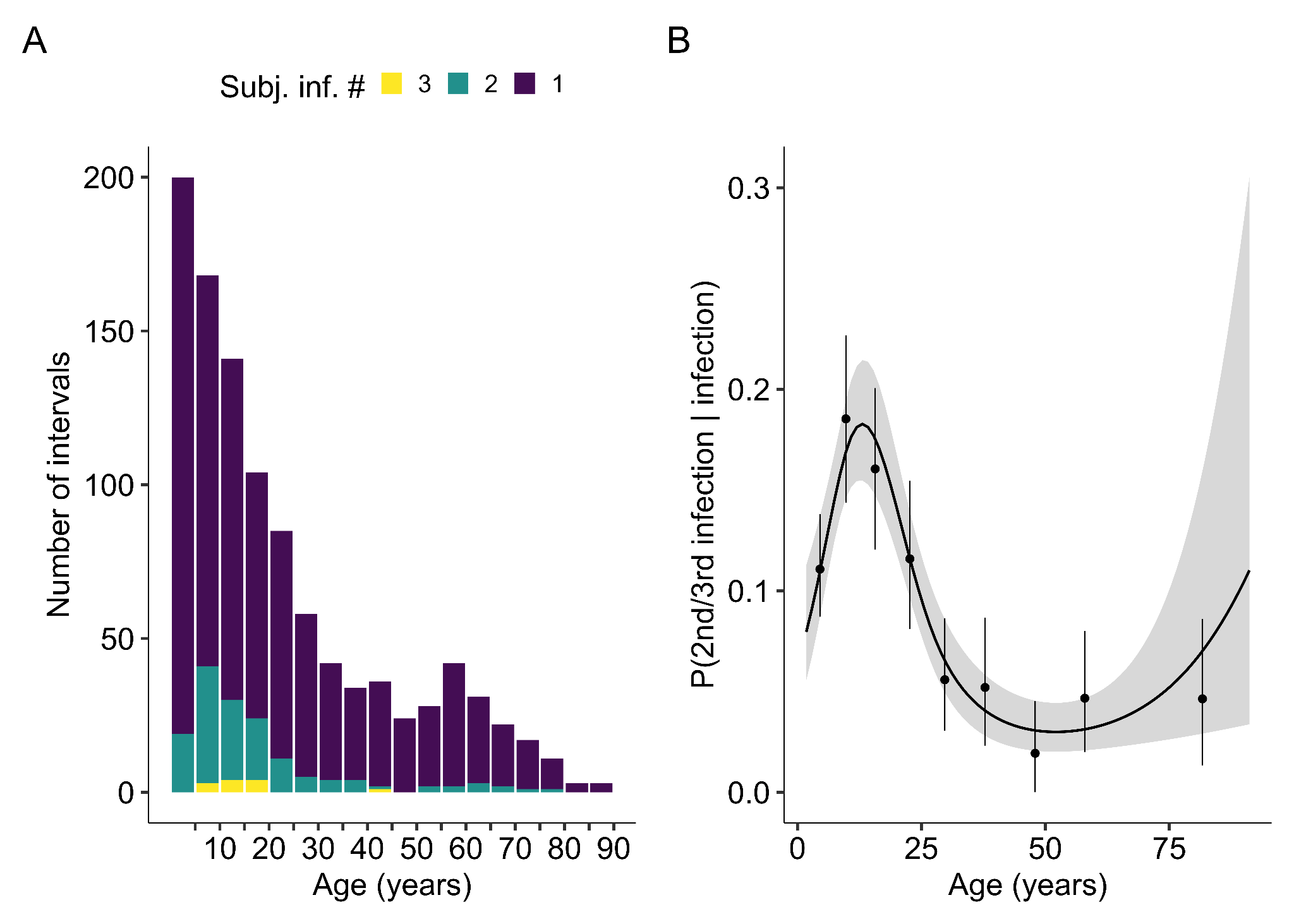

**Figure S3.** (A) Distribution of the number of intervals predicted to have an infection as a function of age at post–interval follow up date. Intervals are color coded by the number of cumulative infections the subject in said interval has had across the study period. (B) Probability of experiencing re-infections during the study by age.

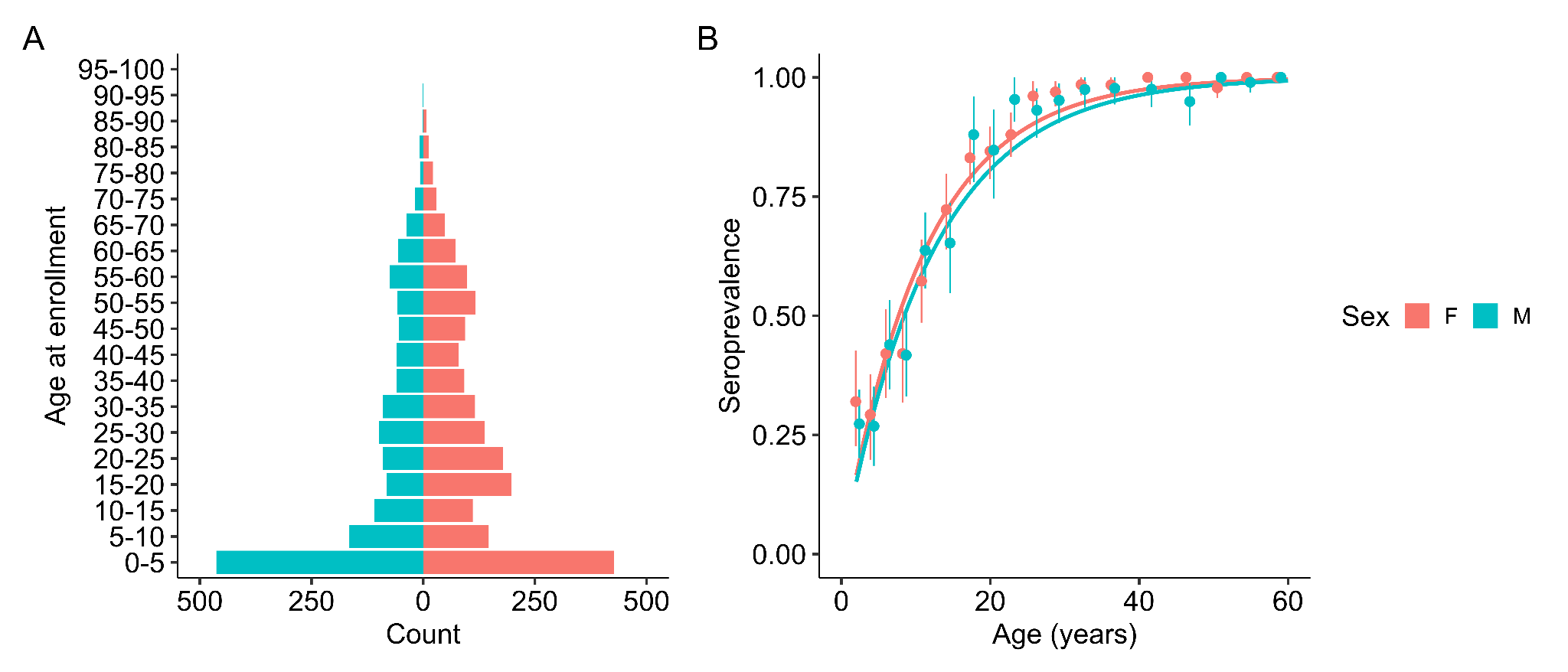

**Figure S4.** (A) Age pyramid in five year bins separated by sex. (B) Seroprevalence curve for subjects enrolled before 2017 in solid line, the analysis was conducted separately by sex. Points are mean seroprevalence in each fifth percentile with newborns removed. Confidence bounds are found using a basic nonparametric bootstrap.

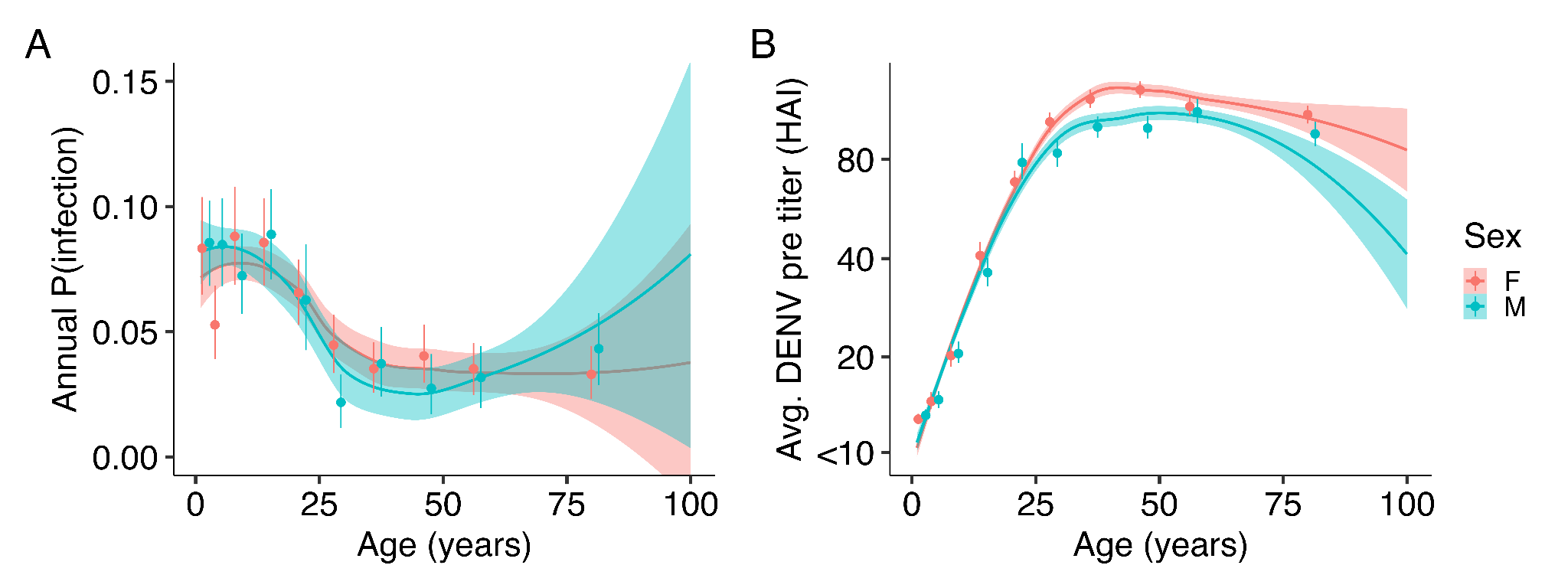

**Figure S5.** (A) Pre-interval DENV titers averaged across all four serotypes and their impact on probability of infection. (B) Relationship between pre-interval DENV titers averaged across all four serotypes and age.

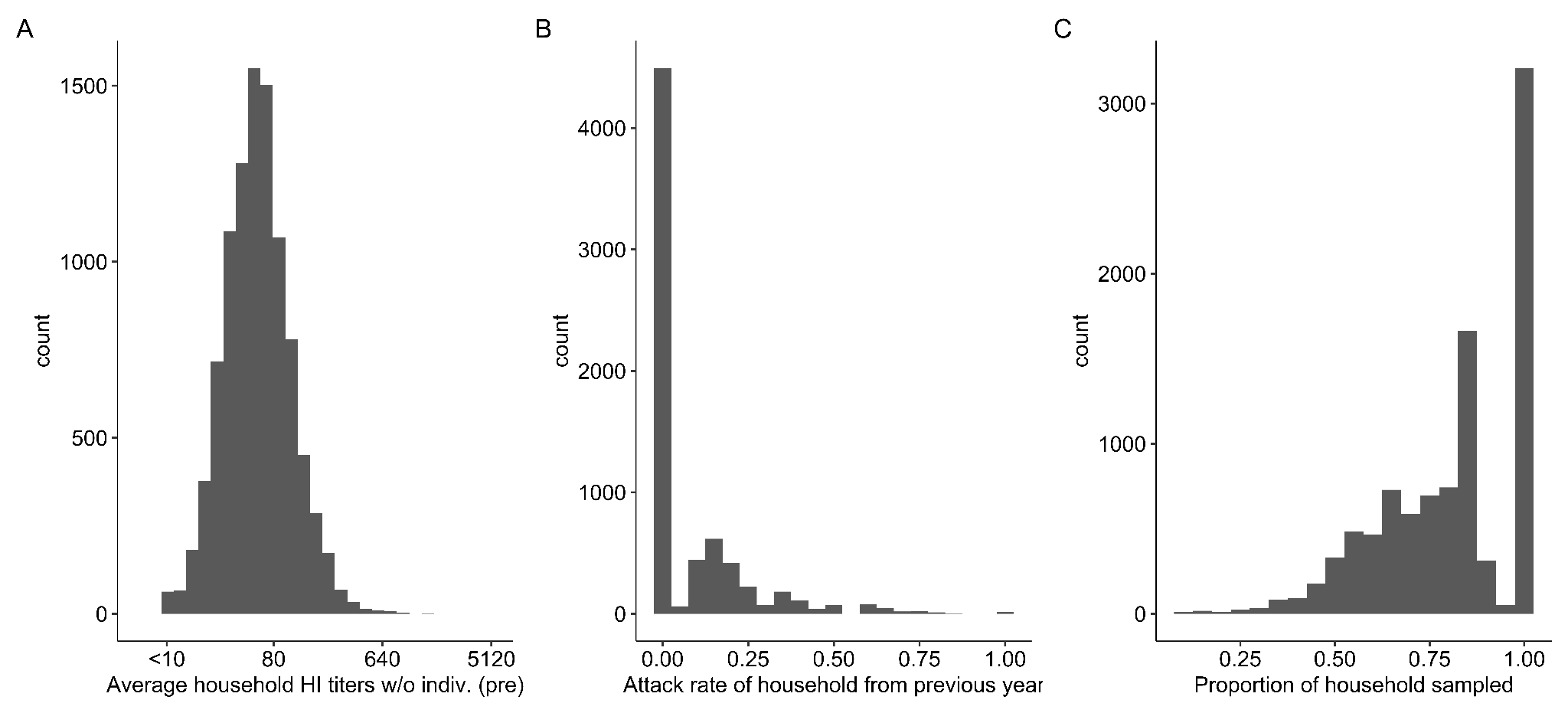

**Figure S6.** Distribution of household related variables including (A) average household HAI titer not including the individual of interest, (B) attack rate of households from the previous year, and (C) the proportion of a household that is sampled.

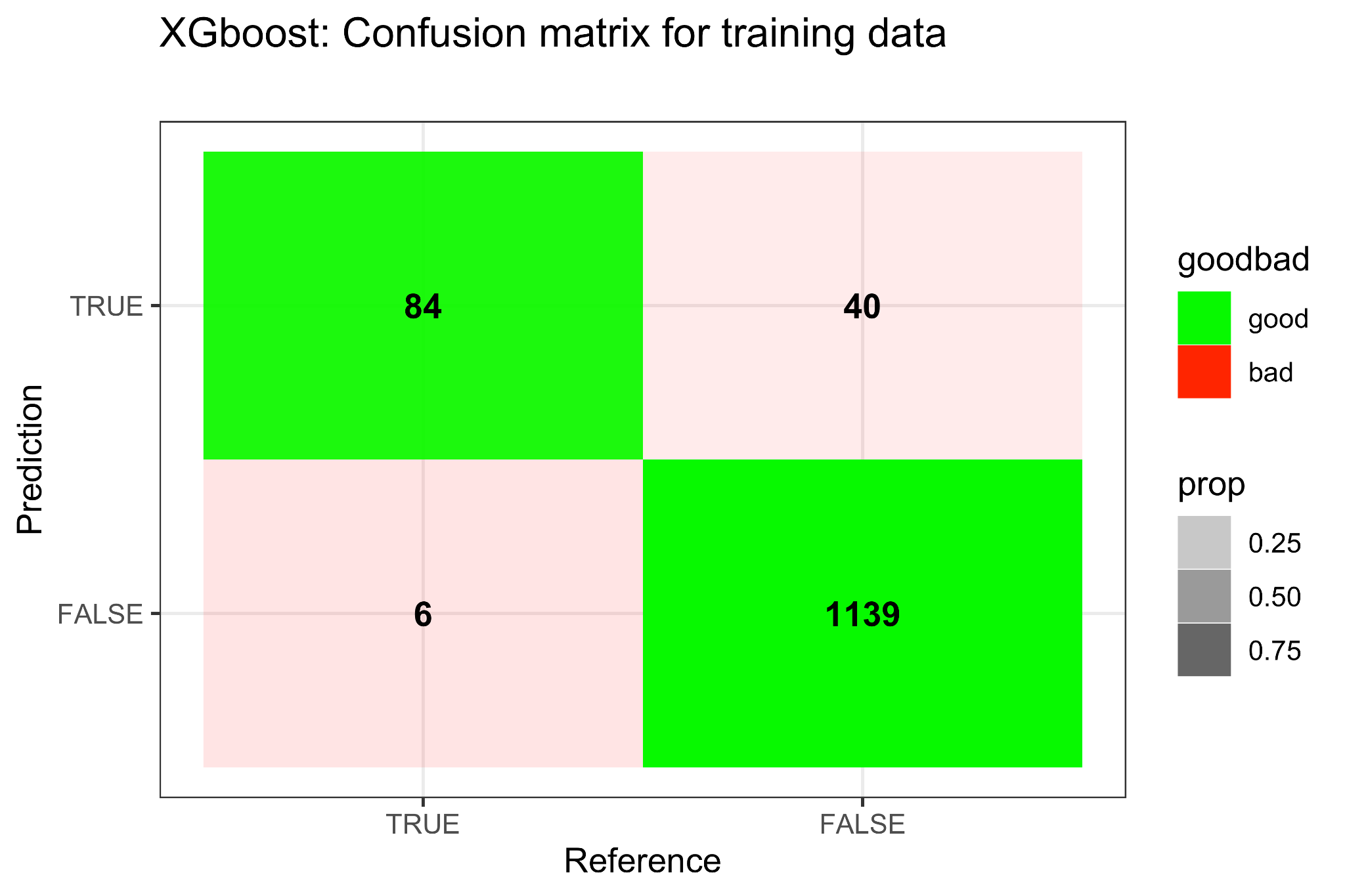

**Figure S7.** Confusion matrix for XGBoost model on training data. Reference values are found as outlined in the Methods section titled Training data while prediction values are found using the approach outlined in the Model fit section of the methods section.

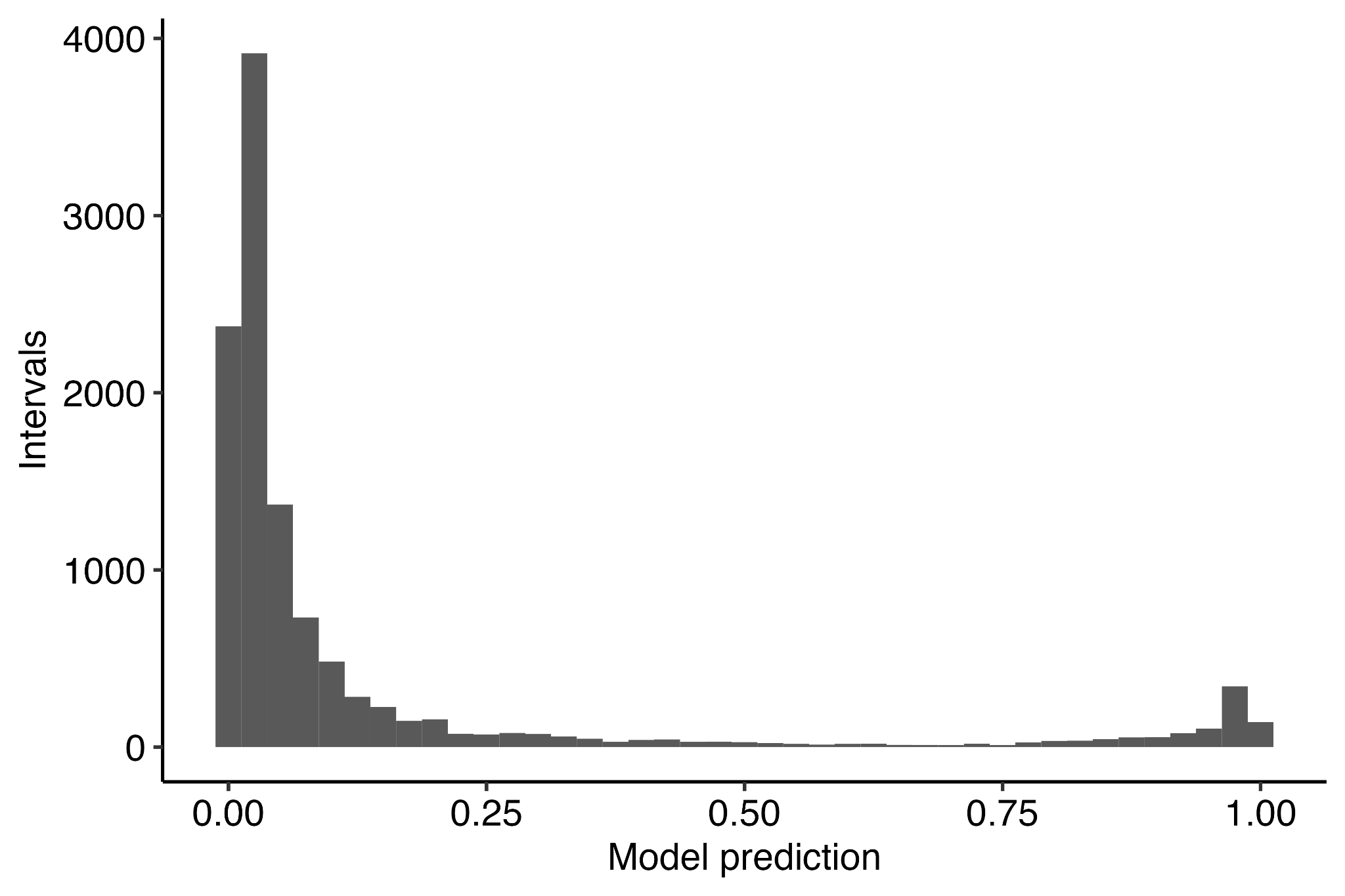

**Figure S8.** Histogram of model predictions for probability of infection between sequential blood draws conducted on the full dataset.

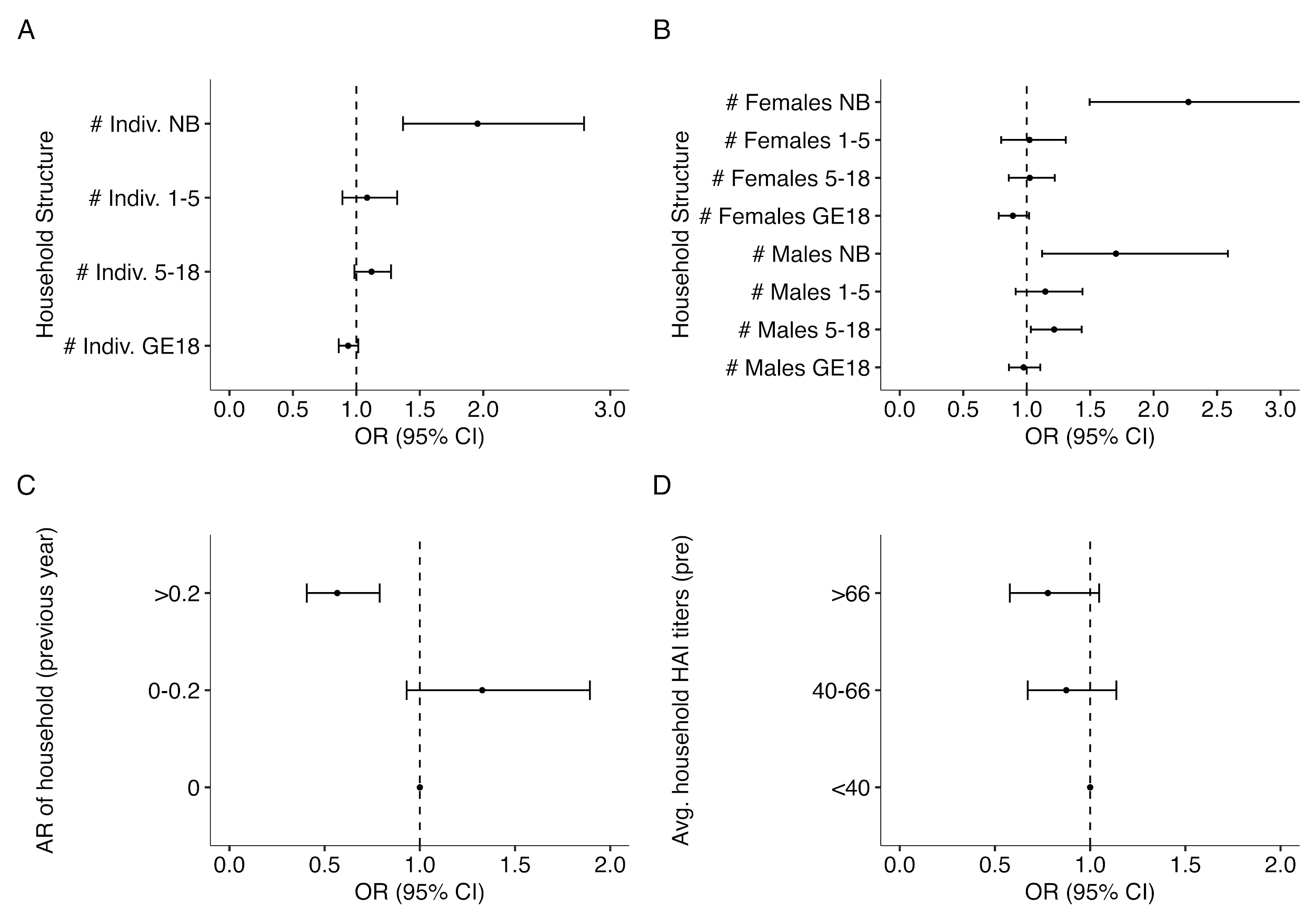

**Figure S9.** Sensitivity analysis on how household composition (A-B), infection history (C) and immunity (D) impact risk of infection on households where more than 80% of samples are recorded (n=6435). (A) Odds ratio for the number of total individuals in various age bins (newborn [NB], from 1-5 years old [LT5], from 5 to 18, and those older than 18 [GE18]) defined at the time of the post-interval sample. (B) Odds ratio for the number of males and females of various age bins (newborn [NB], from 1-5 years old [LT5], from 5 to 18, and those older than 18 [GE18]) defined at the time of the post-interval sample. (C) Previous interval’s attack rate and subsequent odds ratio of infection risk relative to having no infections in the previous interval. (D) Geometric mean of DENV HAI titers for the rest of the household and subsequent odds ratio of infection risk relative to having an average household HAI titer under 40. All models are adjusted for household random effects, individual pre-interval titers, as well as the year and month of post-interval sample.

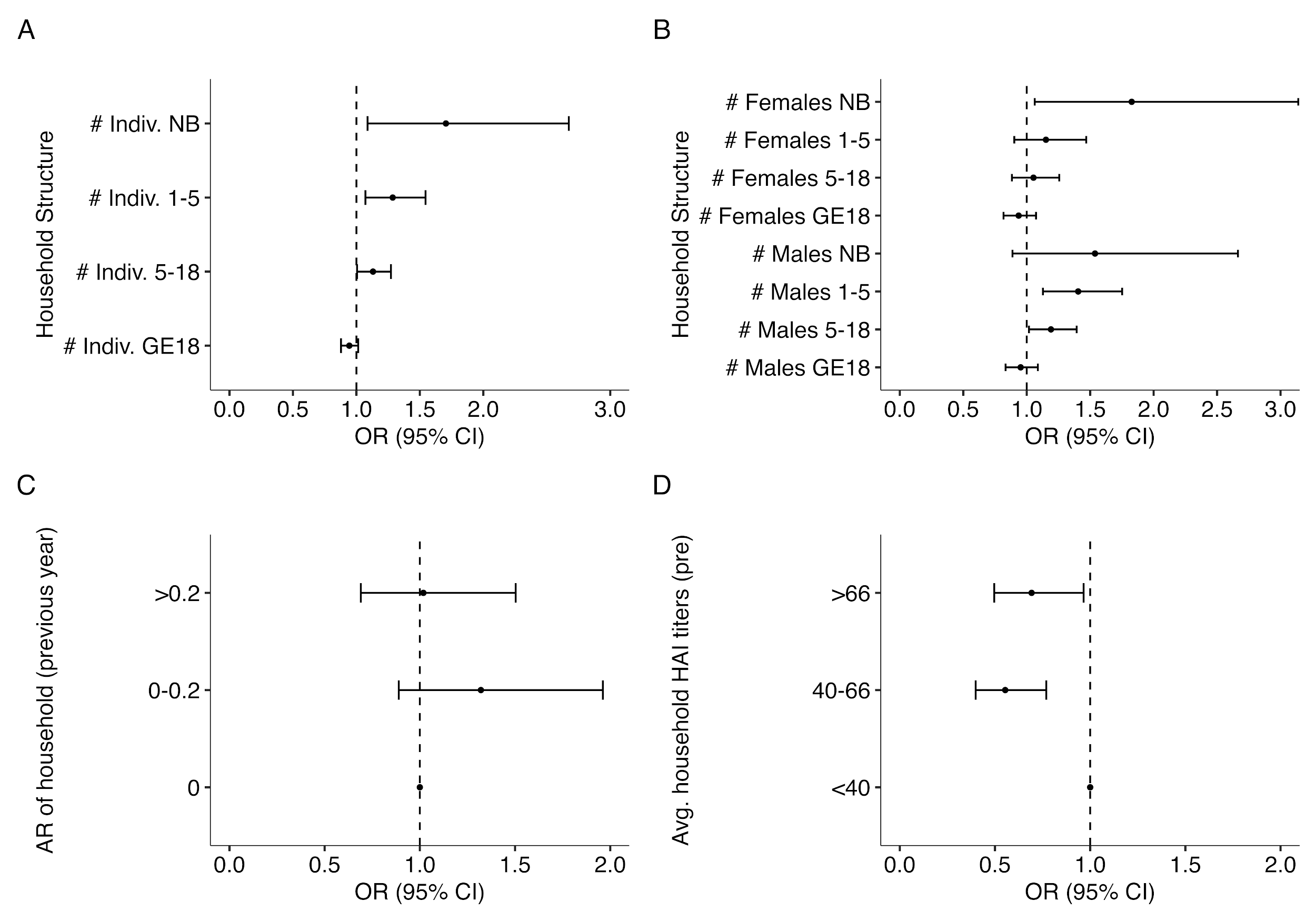

**Figure S10.** Sensitivity analysis on how household composition (A-B), infection history (C) and immunity (D) impact risk of infection on individuals who were seronaive at the start of an interval (n=2066). (A) Odds ratio for the number of total individuals in various age bins (newborn [NB], from 1-5 years old [LT5], from 5 to 18, and those older than 18 [GE18]) defined at the time of the post-interval sample. (B) Odds ratio for the number of males and females of various age bins (newborn [NB], from 1-5 years old [LT5], from 5 to 18, and those older than 18 [GE18]) defined at the time of the post-interval sample. (C) Previous interval’s attack rate and subsequent odds ratio of infection risk relative to having no infections in the previous interval. (D) Geometric mean of DENV HAI titers for the rest of the household and subsequent odds ratio of infection risk relative to having an average household HAI titer under 40. All models are adjusted for household random effects, individual pre-interval titers, as well as the year and month of post-interval sample.

**Table S2.** Odds ratios and adjusted odds ratios for temporal covariates of infection risk. Univariate analyses inform which variables are incorporated in the multivariate analyses. Multivariate analyses were conducted using either just household random effects, both household and individual random effects, or household random effects and incorporating individual immunity.

| **Covariate** | | **Univariate** | **Multivariate**  **House res** | **Multivariate**  **indiv., house res** | **Multivariate house res w/**  **indiv. immunity** |
| --- | --- | --- | --- | --- | --- |
|  |  | **OR**  **(95% CI)** | **aOR**  **(95% CI)** | **aOR**  **(95% CI)** | **aOR**  **(95% CI)** |
| **Temporal level** | | | | | |
| Year | 2015 | REF | REF | REF | REF |
|  | 2016 | 0.56 (0.36 - 0.88) | 0.47 (0.27 - 0.83) | 0.47 (0.26 - 0.82) | 0.48 (0.27 - 0.85) |
|  | 2017 | 0.77 (0.50 - 1.16) | 0.71 (0.42 - 1.20) | 0.70 (0.41 - 1.20) | 0.73 (0.43 - 1.26) |
|  | 2018 | 1.55 (1.03 - 2.32) | 1.21 (0.71 - 2.06) | 1.21 (0.71 - 2.08) | 1.27 (0.74 - 2.19) |
|  | 2019 | 1.58 (1.05 - 2.37) | 1.38 (0.77 - 2.46) | 1.39 (0.78 - 2.50) | 1.51 (0.84 - 2.72) |
|  | 2020 | 1.43 (0.95 - 2.17) | 1.47 (0.77 - 2.79) | 1.49 (0.77 - 2.86) | 1.58 (0.82 - 3.05) |
|  | 2021 | 0.90 (0.54 - 1.51) | 0.58 (0.29 - 1.15) | 0.57 (0.29 - 1.14) | 0.60 (0.30 - 1.20) |
| Month | Jan. | 2.95 (2.21 - 3.94) | 2.13 (1.43 - 3.15) | 2.13 (1.43 - 3.15) | 2.11 (1.40 - 3.17) |
|  | Feb. | 1.80 (1.31 - 2.48) | 1.57 (1.02 - 2.43) | 1.57 (1.02 - 2.43) | 1.58 (1.01 - 2.47) |
|  | Mar. | 1.30 (1.01 - 1.67) | 0.96 (0.69 - 1.34) | 0.96 (0.69 - 1.34) | 0.95 (0.68 - 1.33) |
|  | Apr. | 1.12 (0.85 - 1.47) | 1.05 (0.75 - 1.47) | 1.05 (0.75 - 1.47) | 1.04 (0.74 - 1.47) |
|  | May | REF | REF | REF | REF |
|  | Jun. | 1.37 (1.09 - 1.73) | 1.27 (0.95 - 1.71) | 1.27 (0.95 - 1.71) | 1.26 (0.93 - 1.70) |
|  | Jul. | 1.66 (1.24 - 2.22) | 0.84 (0.53 - 1.33) | 0.84 (0.53 - 1.33) | 0.83 (0.52 - 1.32) |
|  | Aug. | 1.91 (1.42 - 2.58) | 0.90 (0.55 - 1.48) | 0.90 (0.55 - 1.48) | 0.90 (0.54 - 1.49) |
|  | Sep. | 2.17 (1.66 - 2.84) | 1.12 (0.71 - 1.79) | 1.12 (0.71 - 1.79) | 1.18 (0.74 - 1.91) |
|  | Oct. | 2.26 (1.65 - 3.08) | 1.16 (0.70 - 1.93) | 1.16 (0.70 - 1.93) | 1.19 (0.71 - 2.00) |
|  | Nov. | 1.36 (0.83 - 2.22) | 0.48 (0.23 - 1.02) | 0.48 (0.23 - 1.02) | 0.50 (0.23 - 1.07) |
|  | Dec. | NA | NA | NA | NA |

**Table S3.** The following predictors are incorporated into the XGboost model for training and prediction.

| **Predictor** | **Units** |
| --- | --- |
| Individual pre-interval DENV 1-4 and JEV titers | HAI |
| Geometric mean of pre-interval DENV 1-4 titers | HAI |
| Maximum of pre-interval DENV 1-4 titers | HAI |
| Minimum of pre-interval DENV 1-4 titers | HAI |
| Variance of pre-interval DENV 1-4 titers | HAI |
| Individual post-interval DENV 1-4 and JEV titers | HAI |
| Geometric mean of post-interval DENV 1-4 titers | HAI |
| Maximum of post-interval DENV 1-4 titers | HAI |
| Minimum of post-interval DENV 1-4 titers | HAI |
| Variance of post-interval DENV 1-4 titers | HAI |
| Serotype and JEV specific post to pre-interval titers | Ratio |
| Mean of post to pre-interval DENV 1-4 titer ratios | Ratio |
| Maximum of post to pre-interval DENV 1-4 titer ratios | Ratio |
| Minimum of post to pre-interval DENV 1-4 titer ratios | Ratio |
| Variance of post to pre-interval DENV 1-4 titer ratios | Ratio |
| Serotype and JEV specific post minus pre-interval titers | HAI |
| Geometric mean of post minus pre-interval DENV 1-4 titers | HAI |
| Maximum of post minus pre-interval DENV 1-4 titers | HAI |
| Minimum of post minus pre-interval DENV 1-4 titers | HAI |
| Variance of post minus pre-interval DENV 1-4 titers | HAI |
| Sex | NA |
| Time between samples | Days |
| Date of pre-interval sample | Days |
| Date of post-interval sample | Days |
| Age at post-interval sample | Years |
| Age at enrollment | Years |
